# Uses of pathogen detection data to estimate vaccine direct effects in case-control studies

**DOI:** 10.1101/2020.01.15.20017749

**Authors:** Joseph A. Lewnard

**Author notes:** Correspondence to Dr. Joseph Lewnard, Division of Epidemiology and Biostatistics, School of Public Health, University of California, Berkeley, 2121 Berkeley Way, Berkeley, CA 94720. Funding: The author received no specific funding for this article.

## Abstract

The diagnosis of infectious disease syndromes such as fever, diarrhea, and pneumonia is complicated by the potential for shedding or carriage of putatively etiologic pathogens among individuals experiencing symptoms due to other causes. Symptomatic individuals among whom a pathogen is detected, but whose symptoms are caused by other factors, may be misclassified by diagnostic criteria based on pathogen detection. Case-control studies are commonly undertaken to estimate vaccine effectiveness, and present an opportunity to compare pathogen detection among individuals with and without clinically-relevant symptoms. Considering this study context, we derive simple estimators for the direct effects of vaccination on various aspects of host susceptibility. These include protection against acquisition of the pathogen of interest, and protection against progression of this pathogen to disease following acquisition. We assess the impact of test sensitivity on these estimators, and extend our framework to identify a “vaccine probe” estimator for pathogen-specific etiologic fractions. We also derive biases affecting vaccine effectiveness estimates under the test-negative design, an alternative approach using similar data. Our results provide strategies for estimation of pathogen-specific vaccine effectiveness in the absence a diagnostic gold standard. These approaches can inform the design and analysis of studies addressing numerous pathogens and vaccines.

---

Attribution of infectious disease syndromes to a specific microbiological agent often involves detection of this agent, or its genetic material, among individuals experiencing relevant clinical signs and/or symptoms. However, in certain instances, detection of a pathogenic organism from a symptomatic individual does not provide a specific determination of disease etiology. Such is the case for pathogens that may be shed or carried in a subclinical state as part of their natural history. For instance, viruses and bacteria that cause pneumonia may be found in respiratory tract specimens from individuals experiencing acute symptoms as well as individuals without clinically-apparent illness, especially among children (1). Similar circumstances arise in the natural history of pathogens that cause diarrhea (2) and acute febrile illness (3). Because many or most acquisitions of these pathogens do not progress to disease, only a proportion of symptomatic individuals among whom these pathogens are detectable may in fact be experiencing pathogen-attributable illness (4).

This circumstance creates challenges for studies of the effectiveness of vaccines and other pathogen-specific interventions. Whereas such studies typically aim to assess whether an intervention prevents a well-defined endpoint of disease attributable to the pathogen of interest, the absence of a gold-standard diagnostic tool may lead to misclassification of individuals’ outcomes. Recently, large-scale studies of pneumonia (5) and diarrhea (6) etiology among children have employed case-control frameworks, enrolling symptomatic cases and community controls to estimate pathogen-attributable fractions of disease burden. The inferential methods of these studies center on a comparison of the prevalence of each pathogen among symptomatic cases and asymptomatic controls (7–9). While similar case-control frameworks may present an opportunity to correct for misclassification when estimating vaccine effectiveness (VE; (10–14)), such approaches have not been routinely incorporated into real-world studies.

Here we introduce strategies for estimation of VE against pathogen-specific endpoints in case-control studies. As estimands, we consider vaccine direct effect parameters that compare counterfactual outcomes for an individual who does or does not receive vaccination (15). These effects address the biologic reduction in susceptibility conferred by an individual’s response to vaccination, including protection against shedding or carriage of the pathogen, protection against progression of the pathogen to disease (conditioned on acquisition), and the cumulative result of these two forms of protection.

## STUDY DESIGN

### Framework

Consider a case-control study that enrolls cases experiencing a clinical syndrome of interest, which may be preventable by vaccination against one of several causative pathogens. Consider that the study enrolls controls based on clinical criteria unrelated to the pathogen of interest and to vaccination. Such controls may be asymptomatic persons, or those experiencing an “alternative” disease, which does not affect and is not affected by acquisition of the pathogen of interest or vaccination (16). Assume that all or a proportion of individuals enrolled (including both cases and controls) are tested to determine presence or absence of the pathogen of interest.

For the methods described below, we envision a study where there is no *a priori* basis for determining whether a pathogen, once detected, is the true cause of symptoms for an individual patient. We also present simplifications of the approach for contexts where pathogen detection provides a definitive determination of disease etiology.

### Examples

Case-control studies involving the detection of enteropathogens from diarrhea cases and asymptomatic controls, or the detection of respiratory viruses and bacteria in the upper respiratory tract of case with and controls without acute respiratory symptoms, present ideal environments for application of the approaches detailed below. Studies of diarrhea-causing pathogens are a compelling example because the gastrointestinal tract is the site of both colonization and disease for such organisms. Testing may be performed on stool specimens regardless of the presence of symptoms, and the detection of a pathogen in diarrheal stool is not always a clear indication of its etiologic significance (8,17,18). Similar concerns arise with detection of arboviruses, *Rickettsia*, and malaria parasites among individuals experiencing acute febrile illness in endemic settings (3). Although these challenges may be mitigated for certain pathogens by the use of quantitative molecular diagnostic tests (4), such testing remains relatively uncommon in clinical practice.

In studies of pediatric pneumonia, inaccessibility of the site of infection (i.e., the lung) presents a unique challenge where the approaches we consider below may offer further value. The upper respiratory tract (including the oropharynx and nasopharynx) is a readily accessible site for swabbing, but is also the site of colonization or shedding for many pathogens; thus, identification of a pathogen in the upper respiratory tract of a symptomatic individual is not an indication of its etiologic significance. While lower respiratory tract secretions may be desirable because they originate from the site of infection, contamination with organisms from the upper respiratory tract may undermine the benefit of obtaining such specimens (19).

## NOTATION AND THEORETICAL FRAMEWORK

Define *Y*_*i*_ as an indicator of the case (*Y*_*i*_ = 1) or control (*Y*_*i*_ = 0) status of individual *i* with respect to the clinical syndrome of interest. Define *X*_*i*_ as an indicator of the presence (*X*_*i*_ = 1) or absence (*X*_*i*_ = 0) of the pathogen of interest, and define *Z*_*i*_ as an indicator of the individual’s vaccine status as vaccinated (exposed, *Z*_*i*_ = 1) or unvaccinated (unexposed, *Z*_*i*_ = 0; **Table 1**).

**Table 1:** Variables or observations.

| Variable | Definition |
| --- | --- |
| $Y_i$ | Indicator of case ($Y_i = 1$) or control ($Y_i = 0$) status, as defined by clinical symptoms, for individual $i$ |
| $X_i$ | Indicator of pathogen presence ($X_i = 1$) or absence ($X_i = 0$), for individual $i$ |
| $Z_i$ | Indicator of vaccine receipt ($Z_i = 1$) or non-receipt ($Z_i = 0$), for individual $i$ |
| $W_{i,k}$ | Observation from the $k$th confounding variable for individual $i$ |
| $D_i$ | Indicator that a pathogen is detected ($D_i = 1$) or not detected ($D_i = 0$), for individual $i$ |

We next consider epidemiologic parameters describing the prevalence and natural history of the vaccine-preventable pathogen of interest among cases and controls(**Table 2**). Define the prevalence of carriage or shedding of the pathogen in the absence of disease and vaccine-derived immunity as *π* = Pr (*X*_*i*_ = 1|*Y*_*i*_ = 0, *Z*_*i*_ = 0). Define *θ*_*S*_*π* as the prevalence of shedding or carriage of the pathogen of interest among vaccinated persons not experiencing disease, with VE_*S*_ = 1 − *θ*_*S*_ indicating the reduction in prevalence attributable to vaccine effects on pathogen acquisition or clearance. Here we consider prevalence comparisons rather than time-to-event comparisons due to the nature of data available in case-control studies, which would not include longitudinal observations of acquisition and clearance of the pathogen. We address the interpretation of *θ*_*S*_ with respect to vaccine effects on the acquisition and duration of shedding in a later section.

**Table 2:**
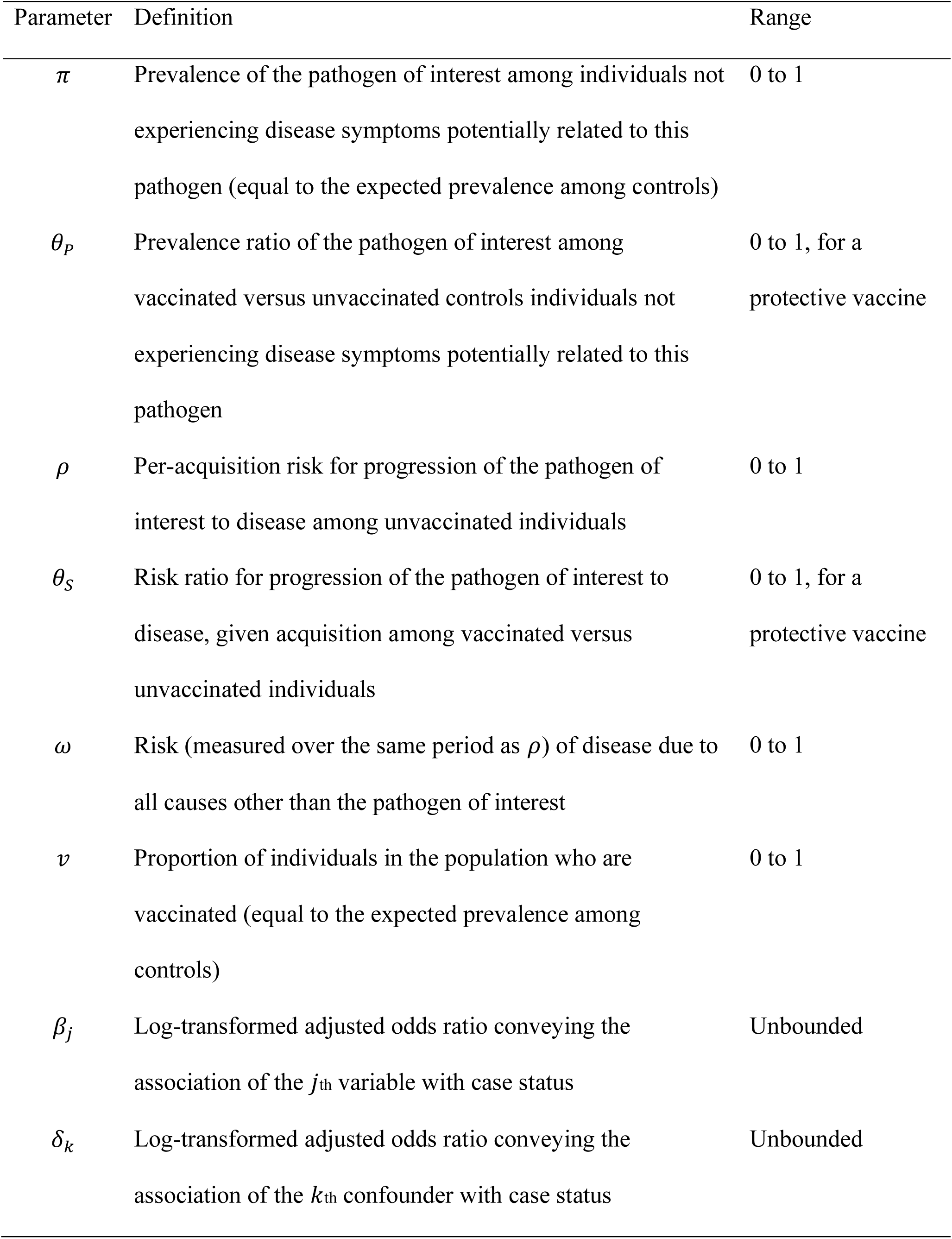
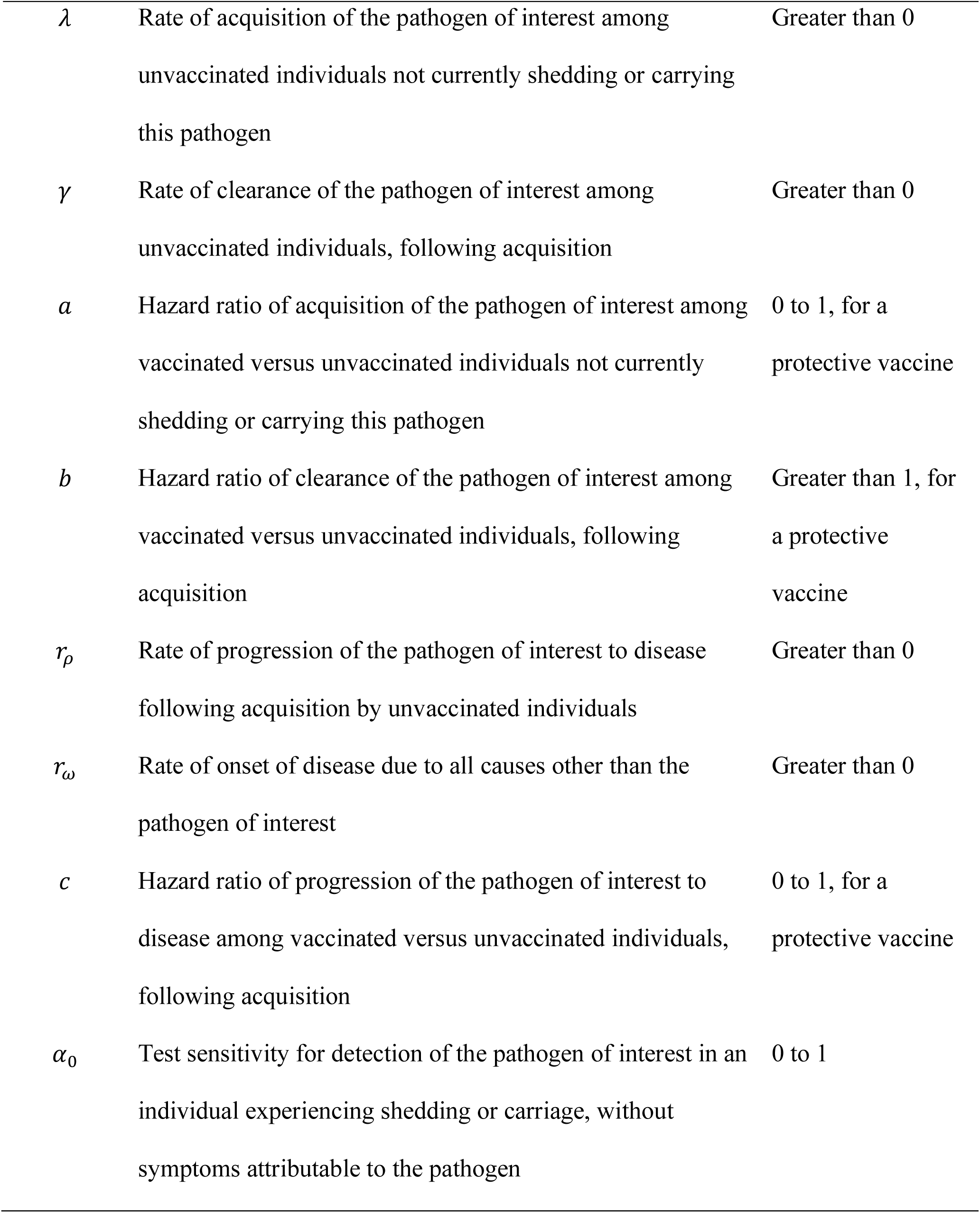
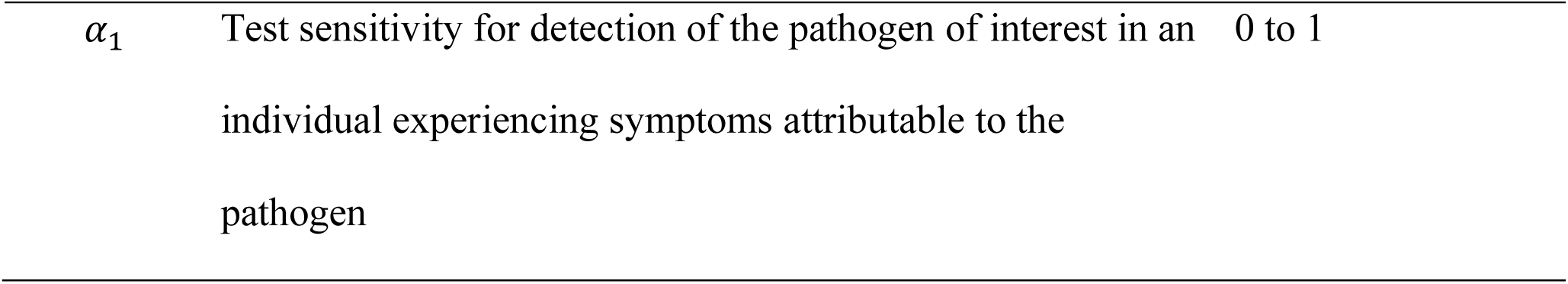
Parameters of the model.

Define *ρ* as the risk for the pathogen of interest, once acquired, to progress to disease in the absence of vaccine-conferred protection. Define *θ*_*P*_ as the relative risk of progression by the pathogen of interest, given vaccination, so that the vaccine direct effect against progression is VE_*P*_ = 1 − *θ*_*P*_. Last, allow *ω* to represent the risk (measured over the same time interval as *ρ*) of disease attributable to all other causes. We assume here that onset of disease due to causes other than the pathogen of interest is unrelated to either prior vaccination or carriage or shedding of the pathogen of interest. We derive *ρ* and *ω* with respect to rate parameters in a later section to assess the relation of *θ*_*P*_ to the hazard ratio of progression of the pathogen of interest to symptomatic disease, given vaccination.

Accounting for disease attributable to the pathogen of interest and all other pathogens, we have

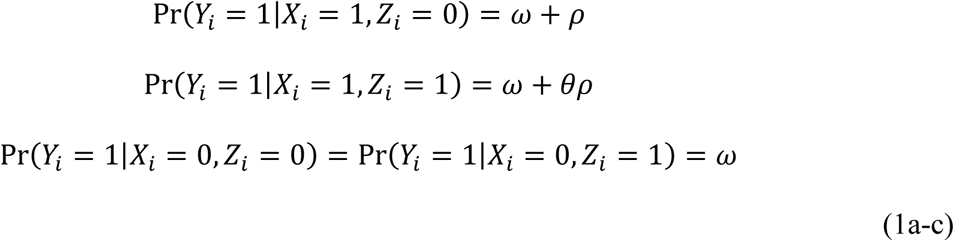

Note that in a case-control study, enrollment of individuals with known outcome status *Y*_*i*_ = 1 or *Y*_*i*_ = 0 hinders direct measurement of Pr(*Y*_*i*_ = *y*|*X*_*i*_ = *x, Z*_*i*_ = *z*). Thus, we consider estimation frameworks below which do not require prospective observation of the probabilities presented in Eq. 1.

## VACCINE DIRECT EFFECT AGAINST SHEDDING OR CARRIAGE OF THE PATHOGEN OF INTEREST (VE_*S*_)

If an individual’s control status is unrelated to risk of shedding the pathogen of interest, the relative risk of detection of the of the pathogen of interest among controls, given vaccination, provides the most straightforward basis for estimating VE_*S*_:

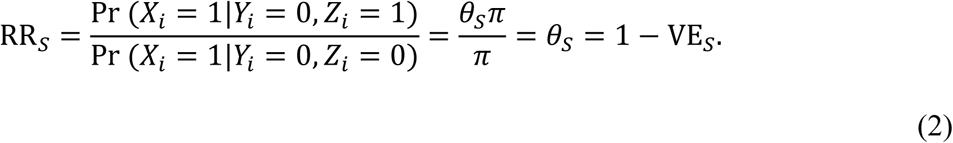

As an alternative, VE_*S*_ can be estimated by comparing the odds of vaccination and pathogen detection among controls against the “null” odds of vaccination within the control group, irrespective of pathogen detection (20). Such an approach may be advantageous when pathogen detection data are available only from a subset of all control individuals from whom vaccination data are available (e.g., in the case of expensive or invasive diagnostic procedures (21,22)), or when favored by other design considerations. This odds ratio (OR) can be expressed as

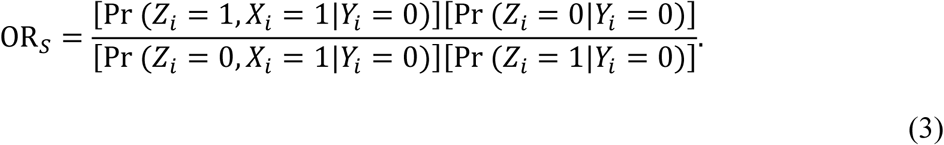

To formalize this intuition, define *v* as the proportion of individuals receiving vaccination in the population; provided vaccination is independent of control status, we have Pr(*Z*_*i*_ = 1|*Y*_*i*_ = 0) = *v*. From the assumption that control status is independent of vaccination and shedding of the pathogen of interest,

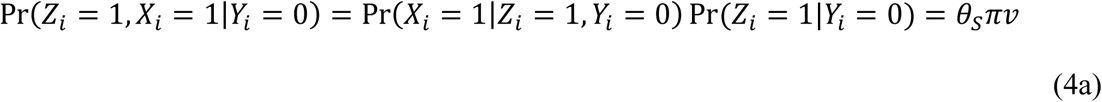

and

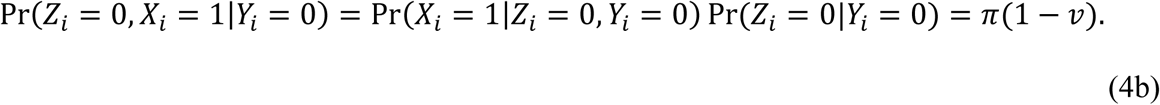

Substituting into Eq. 3,

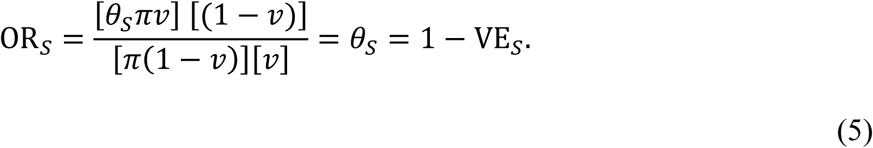

## VACCINE DIRECT EFFECT AGAINST PROGRESSION OF THE PATHOGEN OF INTEREST TO DISEASE FOLLOWING ACQUISITION (VE_*P*_)

### Estimator

Here we address the vaccine-attributable reduction in susceptibility to disease, given a pathogen has overcome a host’s vaccine-derived immunity against acquisition of the carriage or shedding state. To estimate this effect, consider first the OR of detection of the pathogen of interest, given symptoms. We expect this OR to exceed one if acquisition of the pathogen increases individuals’ risk of experiencing symptoms (i.e., *ρ* > 0; (7,8)). Among the unvaccinated,

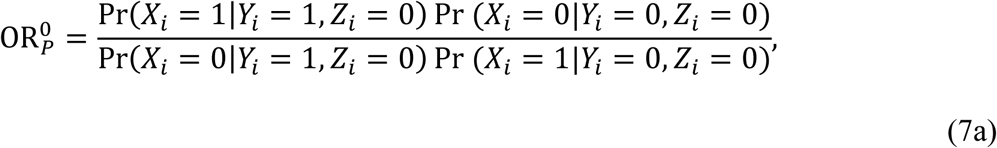

while among the vaccinated,

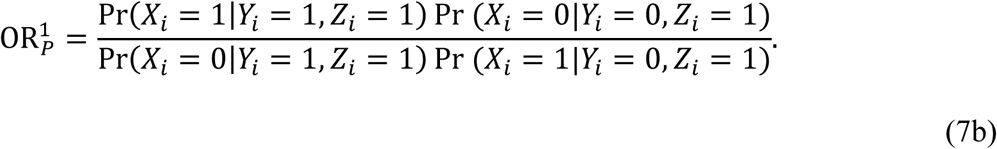

We may expect that 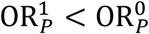 if vaccination confers protection against progression of the pathogen from shedding to disease, thereby attenuating the association between pathogen detection and symptoms among the vaccinated. Of the terms appearing in Eq. 7, we have defined Pr(*X*_*i*_ = 1|*Y*_*i*_ = 0, *Z*_*i*_ = 1) and Pr(*X*_*i*_ = 1|*Y*_*i*_ = 0, *Z*_*i*_ = 0) above in Eq. 2. From the progression parameters *ρ, ω*, and *θ*_*P*_, we may further define

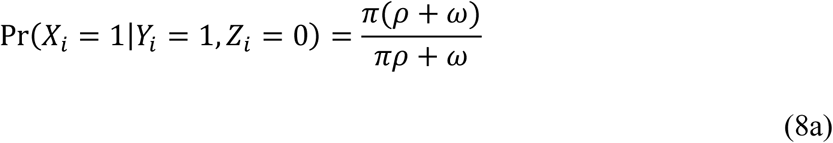

and

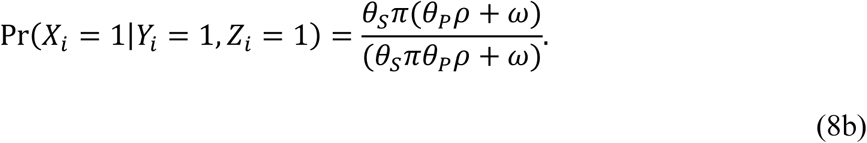

As the complements of Eqs. 8a and 8b, respectively,

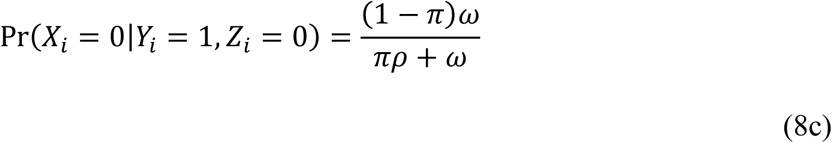

and

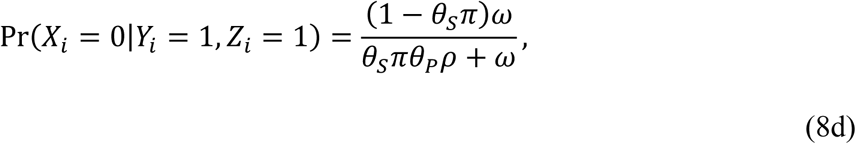

Substituting terms from Eqs. 2 and 8 into Eq. 7, we have

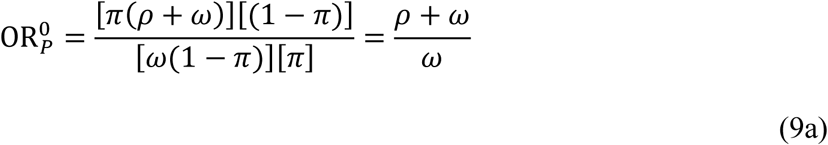

and

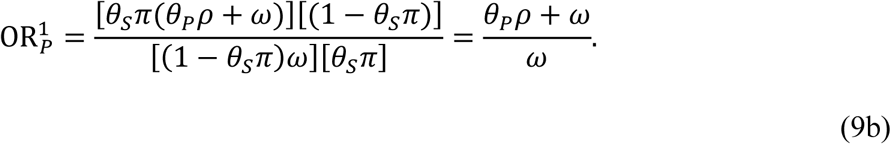

Here, 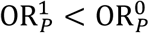 for *θ*_*P*_ < 1 and *ρ* > 0, *ω* > 0. Rearranging the terms from Eq. 9 reveals 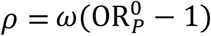 and 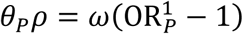. Substituting for *θ*_*P*_,

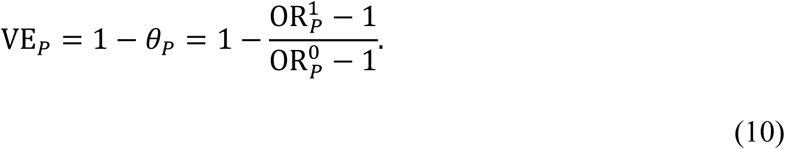

Thus, we may estimate the vaccine direct effect against progression of the targeted pathogen from acquisition to disease by comparing the association of detection of the pathogen with symptoms, among vaccinated and unvaccinated individuals.

### Estimation considerations

Estimation of the vaccine direct effect against progression requires division of the parameters 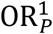 and 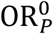. When these quantities are estimated independently, variance of the ratio 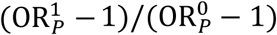 may be overstated due to the failure to account for the covariance of these terms; further, because we envision an observational study design, it may be necessary to control for confounding factors that independently predict individuals’ likelihood of pathogen detection, vaccination, and disease. These concerns may be alleviated by estimating 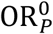 and 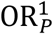 via the logistic regression model

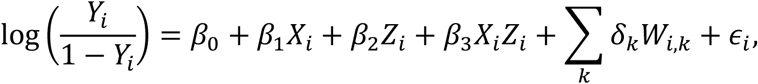

where *ϵ*_*i*_∼N(0, *σ*^2^). Here, we may interpret 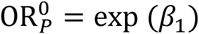 and 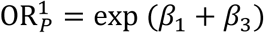 as adjusted estimates of the association between presence of the pathogen and disease risk. The parameters *δ*_*k*_ convey the effects of confounding factors *W*_*k*_, and *β*_2_ conveys any difference in disease risk among the vaccinated and unvaccinated not mediated by protection against the pathogen of interest. Under this framework, we may express the vaccine direct effect against progression in terms of the regression parameters

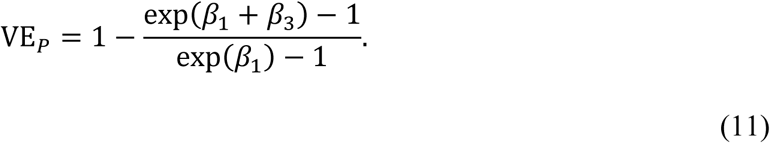

## VACCINE DIRECT EFFECT AGAINST DISEASE ATTRIBUTABLE TO THE PATHOGEN OF INTEREST (VE_*D*_)

We last consider the cumulative extent of vaccine-conferred protection resulting from prevention of shedding or carriage of the pathogen of interest, and prevention of progression to disease in the event of breakthrough acquisition (VE_*D*_). From the joint probabilities of pathogen acquisition and progression among the vaccinated and unvaccinated,

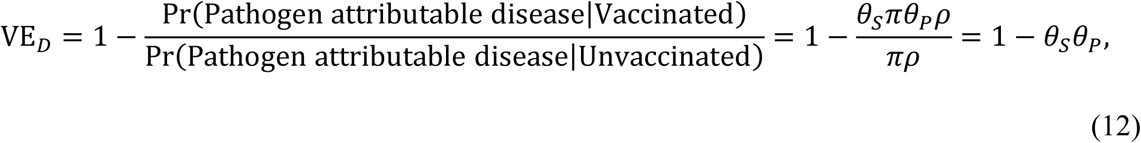

which may thus be estimated as

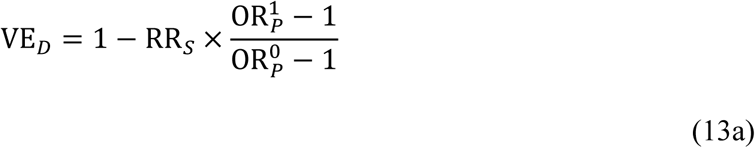

or

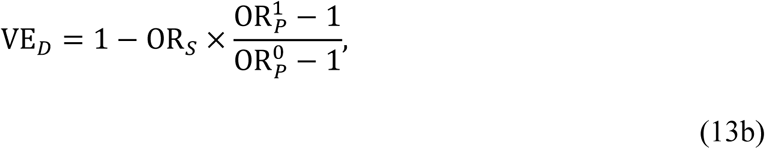

as defined above.

## INTERPRETATION OF RISKS AND RATES

### Pathogen shedding or carriage parameters

Interpretation of pathogen prevalence measures presents a challenge in studies of VE against endpoints such as asymptomatic shedding or carriage of a pathogen due to the dynamic nature of transmission (15,23,24). Here we relate *θ*_*S*_ = 1 − VE_*S*_ to vaccine direct effects on the rates of pathogen acquisition and clearance.

Define *λ* and *γ* as the rates at which unvaccinated individuals acquire and clear the pathogen of interest, respectively. Taking *π* to indicate the equilibrium prevalence of shedding or colonization, with *π* > 0, we have *λ*(1 − *π*) = *γπ* and *π* = *λ*/(*λ* + *γ*). Further defining *a* and *b* as the relative rates of acquisition and clearance of the pathogen, given vaccination, we have *aλ*(1 − *θ*_*S*_*π*) = *bγθ*_*S*_*π*. Substituting for *π*, we obtain

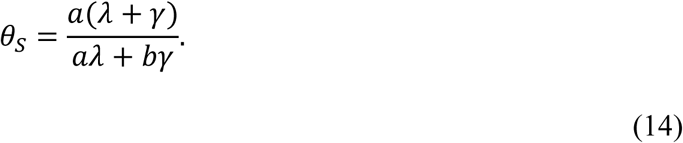

If the rate of clearance of the pathogen far exceeds the rate of acquisition (*γ* ≫ *λ*), we have 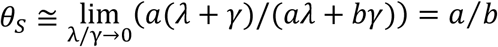. Thus, the prevalence ratio *θ*_*S*_ among controls (as estimated by RR_*S*_ or OR_*S*_) may be interpretable as the hazard ratio of acquisition if *b* = 1, or as the inverse of the hazard ratio of clearance if *a* = 1.

### Disease progression parameters

We may relate the risk parameters *ρ, ω*, and *θ*_*P*_ to epidemiologic rates under a similar intuition. Define *r*_*ρ*_ as the rate of progression of the pathogen of interest to disease, following acquisition by an unvaccinated individual, and define *r*_*ω*_ as the rate of onset of disease due to all other causes. Here we may use a competing risks framework to define *ρ* and *ω* as the probability that disease due to each cause precedes disease due to the other cause, or clearance of the pathogen. Defining *f*(*t*|*r*) as the density function for an event with rate *r* occurring at time *t*, we have

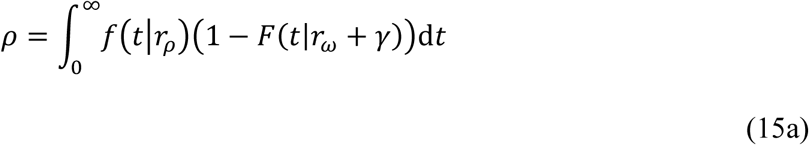

and

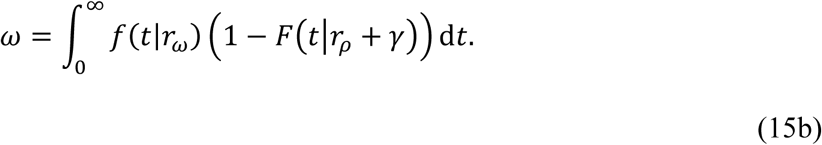

Next defining *c* as the hazard ratio of progression of the pathogen of interest to disease following acquisition, given vaccination, we have

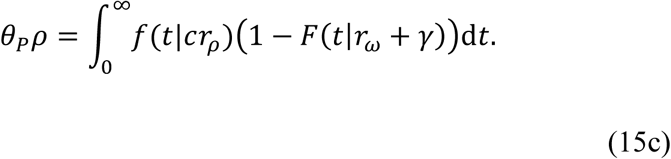

Assuming exponentially-distributed onset times, we have *ρ* = *r*_*ρ*_/(*r*_*ρ*_ + *r*_*ω*_ + *γ*), *ω* = *r*_*ω*_ /(*r*_*ρ*_ + *r*_*ω*_ + *γ*), and *θ*_*P*_*ρ* = *cr*_*ρ*_/(*cr*_*ρ*_ + *r*_*ω*_ + *γ*), so that

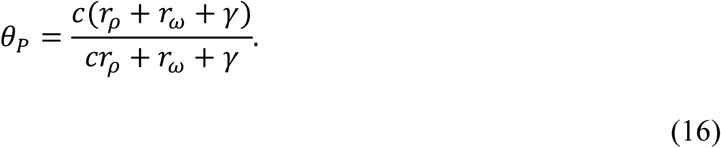

If the rate of progression of the pathogen of interest is well below the rate of disease onset due to all other causes (*r*_*ω*_ ≫ *r*_*ρ*_), or the rate of clearance of the pathogen (*γ* ≫ *r*_*ρ*_), we have 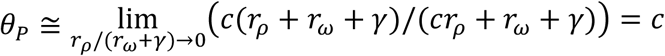. Thus, for diseases that are uncommon, OR_*P*_ = *θ*_*P*_ may be interpretable as the hazard ratio of disease progression, following acquisition, for a vaccinated versus unvaccinated individual.

## STUDIES WITH HIGH DIAGNOSTIC SPECIFICITY IN DISEASE

The framework we introduce above also has value for estimating vaccine-conferred protection against progression to endpoints where the distinction between true disease and subclinical shedding or carriage is unambiguous. For instance, detection of commensal bacteria in ordinarily sterile body fluids such as the bloodstream or cerebrospinal fluid is a defining characteristic of invasive infections involving these organisms. Detection of respiratory viruses in the bloodstream may similarly provide a marker of very severe disease attributable to these pathogens (25–28).

In the event that we may exclude alternative etiologies among cases in whom the pathogen is detected, Eqs. 8a and 8b may be rewritten as

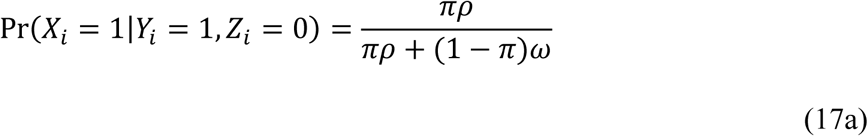

and

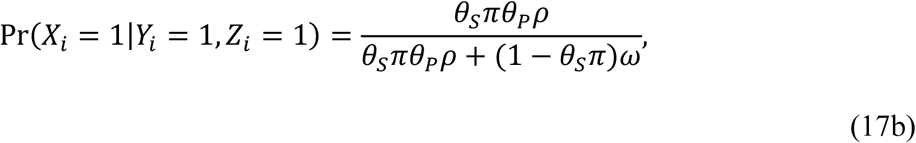

respectively. Here, we subtract *πω*(1 − *v*) and *θ*_*S*_*πωv* from our earlier definitions of Pr(*Y*_*i*_ = 1, *Z*_*i*_ = 0) and Pr(*Y*_*i*_ = 1, *Z*_*i*_ = 1), respectively, because episodes of disease not attributable to the pathogen of interest among cases shedding or carrying this pathogen are not observed. Here,

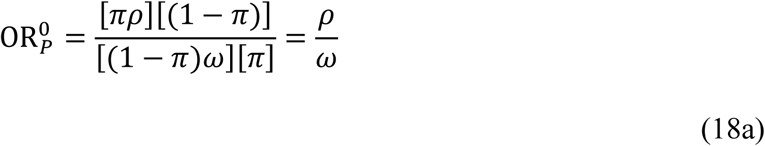

and

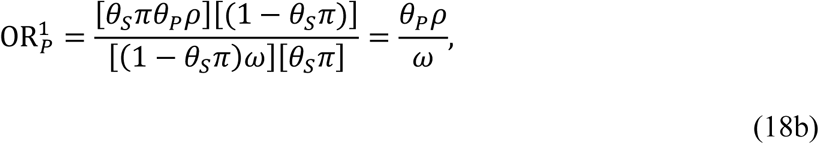

so that

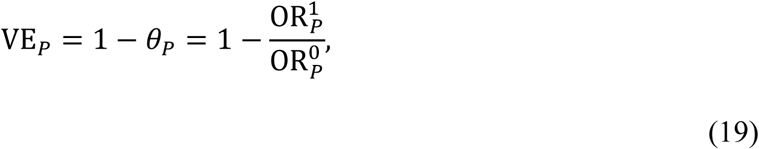

in contrast to Eq.10.

Additionally, this special case allows VE_*D*_ to be estimated directly by comparing the odds of prior vaccination among cases with the pathogen detected versus controls among whom the pathogen is not detected. We show Pr(*Z*_*i*_ = *z*|*X*_*i*_ = 1, *Y*_*i*_ = 0) in Eq. 4. Similarly,

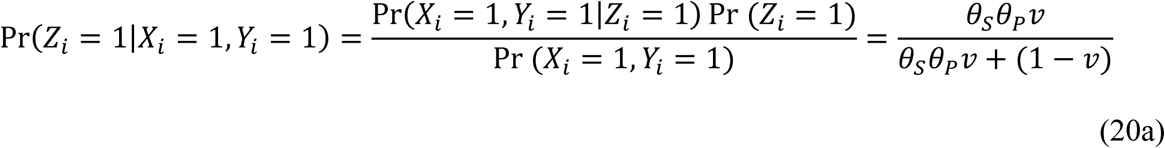

and its complement

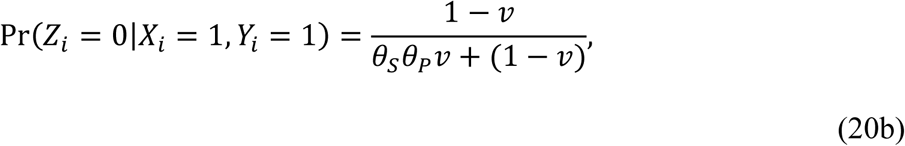

under the circumstance that the pathogen, if detected in a symptomatic individual, is the true cause of disease. Taken together,

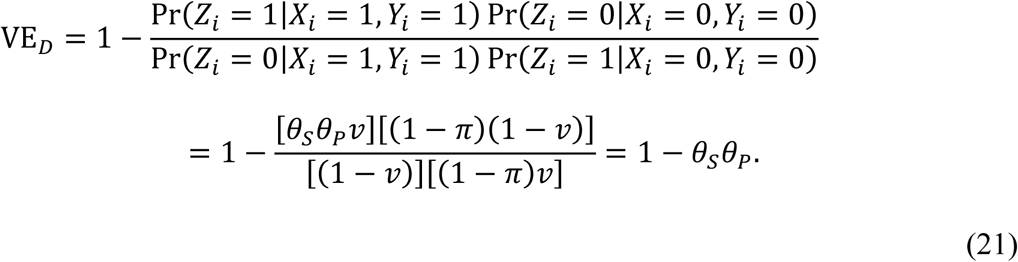

## BIAS UNDER IMPERFECT TEST SENSITIVITY

For certain conditions, sensitivity of pathogen detection approaches may differ for individuals experiencing pathogen-attributable disease versus subclinical shedding or colonization. For instance, higher pathogen load among individuals experiencing disease may improve the probability of pathogen detection among true disease cases via certain diagnostic assays (4,29).

Alternatively, for conditions such as pneumonia involving infection at difficult-to-sample anatomical sites, shedding or carriage at the sampled site (e.g. oronasopharynx) may cease before the clearance of the pathogen from the site of infection (30–32).

Take *D*_*i*_ = 1 and *D*_*i*_ = 0 to indicate detection or non-detection of the pathogen of interest. To account for test sensitivity in the formulations introduced above, we may replace *X*_*i*_ with *D*_*i*_ in our previous equations, so that

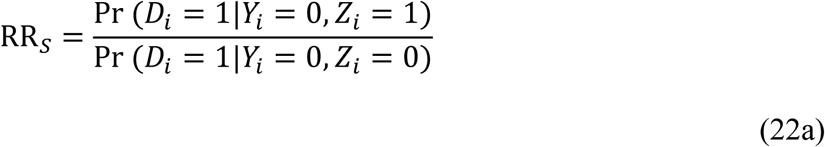

and

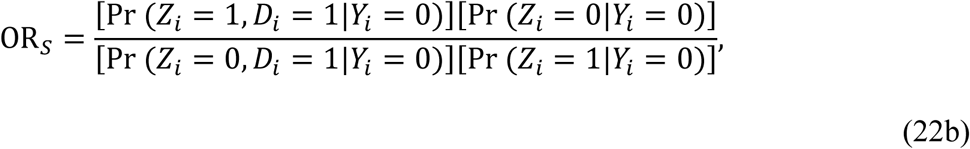

for estimation of VE_*S*_, while

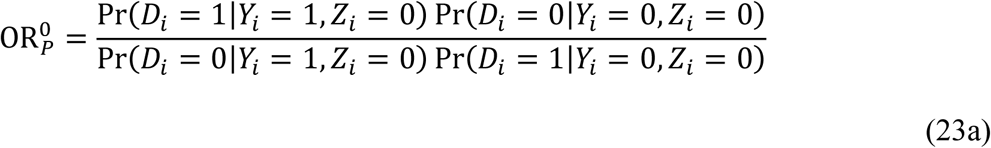

and

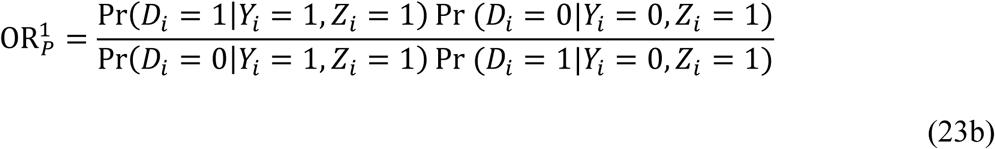

for estimation of VE_*D*_.

Define *α*_1_ and *α*_0_ as the probability of detecting the pathogen of interest, given it is present, for individuals who are and are not experiencing disease attributable to this pathogen. The expected prevalence of pathogen detection among those without symptoms is thus Pr(*D*_*i*_ = 1|*Y*_*i*_ = 0) = *α*_0_ Pr(*X*_*i*_ = 1|*Y*_*i*_ = 0). Substituting into Eq. 17 reveals no effect on RR_*S*_ and OR_*S*_, as *α*_0_ cancels in the numerator and denominator.

Next considering 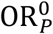, we have

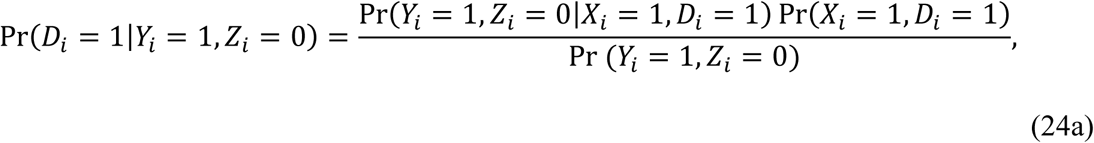

since Pr(*X*_*i*_ = 0, *D*_*i*_ = 1) = 0. Based on the parameter definitions above,

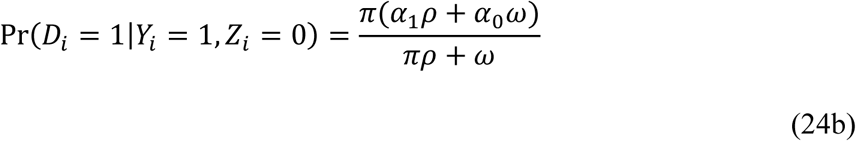

and

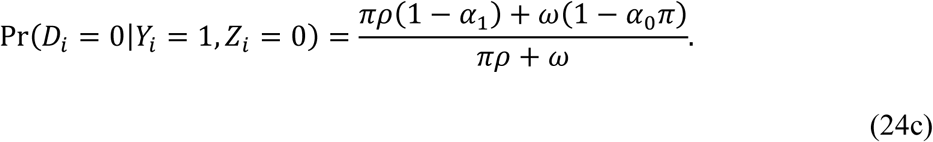

Similarly, among the vaccinated,

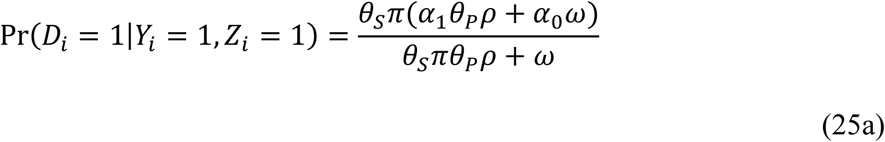

and

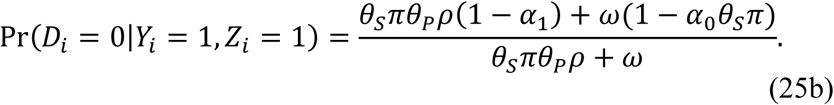

Substituting into Eq. 18, we have

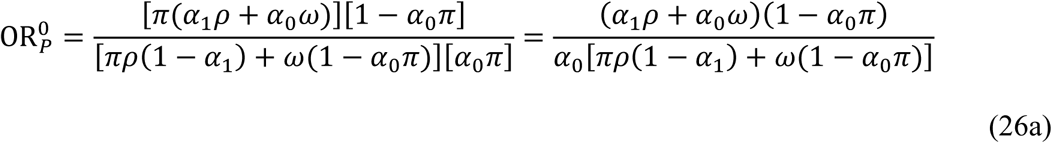

and

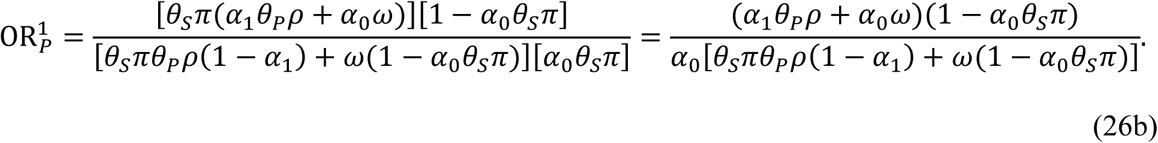

Here, 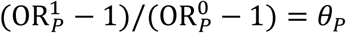 if *α*_1_ = 1; otherwise, 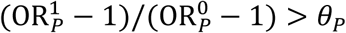 for 0 < *θ*_*P*_ < 1, over the ranges of all other parameters listed in **Table 2**. Thus, if vaccination confers protection against disease progression, imperfect sensitivity affecting pathogen detection among symptomatic cases is expected to lead to underestimation of the true effect.

We illustrate the magnitude of this bias in **Figure 1** under various parameterizations. The extent of bias increases with higher prevalence of the pathogen among persons without symptoms (*π*), with higher rates of progression of the pathogen to disease (*ρ*/*ω*), and with lower sensitivity for pathogen detection in disease (*α*_1_). Interestingly, bias also worsens as *α*_0_ approaches 1.

**Figure 1:**
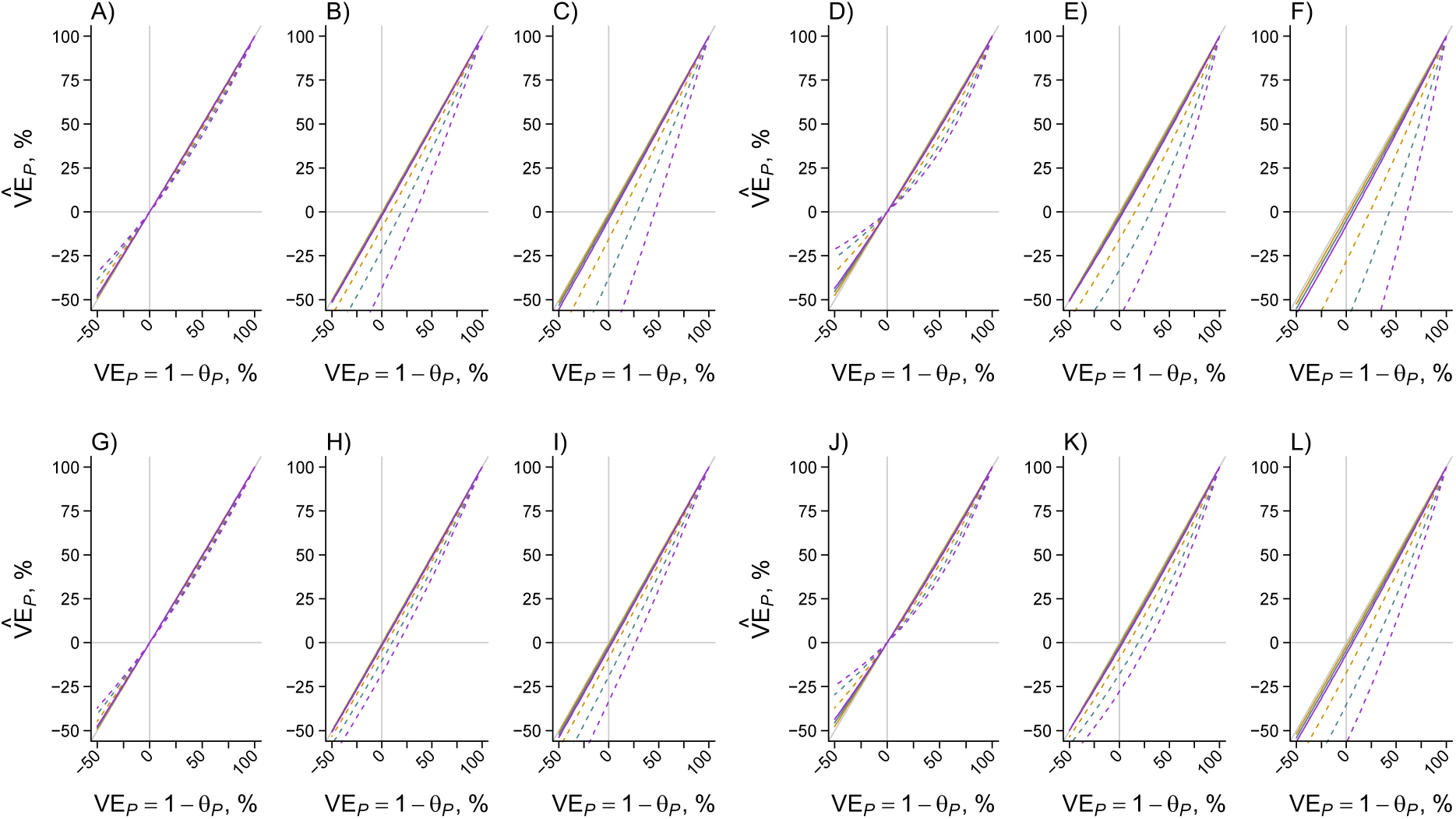
Effect of imperfect test sensitivity on estimates of vaccine-conferred protection against progression of the pathogen to disease. Using the expressions for OR^0^ and OR^1^ in Eq. 26, we plot the estimated effect of vaccination on risk of progression of the pathogen of interest, given acquisition 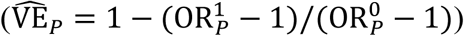. The x-axis indicates the true vaccine effect against progression; departures from the 1:1 diagonal (grey) line indicate bias. Gold, blue, and violet lines correspond to estimates assuming 90%, 80%, and 70% test sensitivity, respectively, for detecting the pathogen of interest among individuals experiencing disease due to this pathogen (gold, *α*_1_ = 0.9; blue, *α*_1_ = 0.8; violet, *α*_1_ = 0.7). Solid and dashed lines correspond to estimates assuming 10% and 50% prevalence, respectively, of the pathogen of interest among unvaccinated individuals without symptoms (solid, *π* = 0.1; dashed, *π* = 0.5). The top row (panels A-F) considers a diagnostic test with 100% sensitivity for detection of the pathogen when it is not a cause of symptoms (*α*_0_ = 1), while the bottom row (panels G-L) considers a test with 70% sensitivity for detecting the pathogen when it is not a cause of symptoms (*α*_0_ = 0.7). Within each column, the left three panels (panels A-C and panels G-I) assume acquisition of the pathogen doubles individuals’ risk of disease (*ρ*/*ω* = 1), while the right three panels (panels D-F and panels J-L) assume acquisition of the pathogen results in a four-fold increase in individuals’ risk of disease (*ρ*/*ω* = 3). Each grouping of three panels presents estimates corresponding to increasing levels of vaccine-conferred protection against shedding or colonization (VE_S_). Left panels (panels A, D, G, J) assume 0% protection (*θ*_*S*_ = 1); center panels (panels B, E, H, K) assume 25% protection (*θ*_*S*_ = 0.75); and right panels (C, F, I, L) assume 50% protection (*θ*_*S*_ = 0.5). Grey lines plotted at VE_*P*_ = 0% and 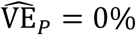 indicate where the true value and estimated value, respectively, cross between indicating increased and decreased risk of disease, given acquisition, as a consequence of vaccination.

The nature and extent of bias further depends upon whether vaccination confers protection against pathogen shedding or carriage (*θ*_*S*_). With *θ*_*S*_ = 1 (i.e., VE_*S*_ = 0), the estimate 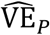 converges to the true value when VE_*P*_ = 0% or VE_*P*_ = 100%, and is sign-unbiased, such that 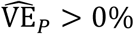 when VE_*P*_ > 0%, and 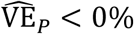 when VE_*P*_ < 0%. In contrast, the estimate may be sign-biased when vaccination confers protection against shedding (i.e., 0 < *θ*_*S*_ < 1); here 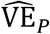 might suggest increased risk of disease progression among vaccinated individuals who acquire the pathogen, when in fact vaccination protects against disease progression. However, this circumstance is expected only under extreme scenarios, where shedding or carriage is very prevalent among asymptomatic individuals, test sensitivity is very high in the context of subclinical shedding or carriage and low among those experiencing disease, and true protection against disease progression is very weak.

## BIAS UNDER TEST-NEGATIVE DESIGN APPROACHES

### Protection against disease

The test-negative design (TND) has come into routine use in studies of VE (33). These studies estimate vaccine direct effects by comparing the odds of prior vaccination among individuals experiencing clinically-apparent illness who test positive or negative for a pathogen of interest 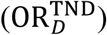. Case-control studies such as we consider here may resemble TND studies in that receipt of a test is independent of the likelihood of detecting the pathogen of interest (14,34,35). It has been previously recognized that shedding or carriage of a pathogen among individuals experiencing symptoms due to other causes may influence TND estimates, although the extent of resulting bias is not well understood (10,12,14). Here, we derive the mathematical form of this bias using the framework described above.

Studies using the TND aim to estimate the vaccine direct effect against disease attributable to the pathogen of interest according to

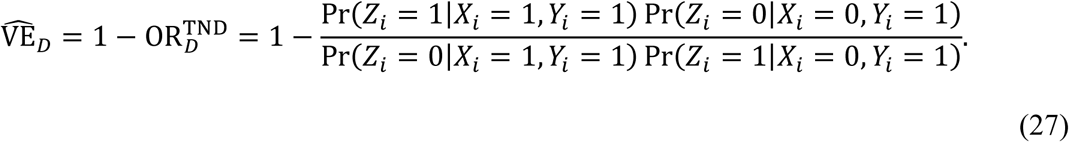

Here we may define

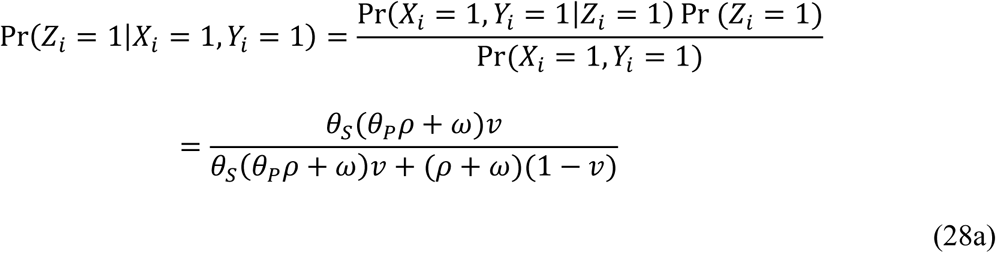

and

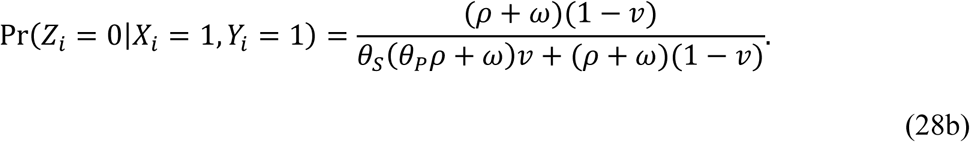

Similarly, we have

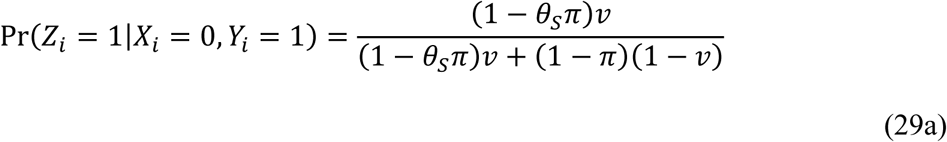

and

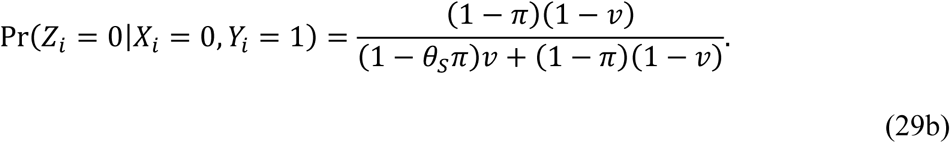

Substituting Eqs. 28 and 29 into Eq. 27, we obtain

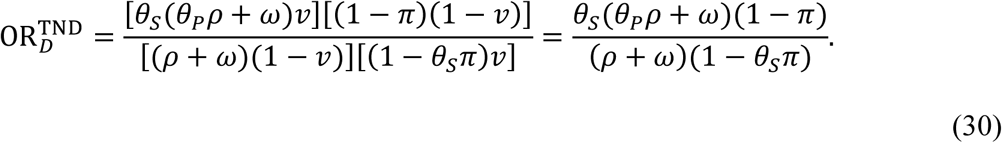

This estimator approaches the true vaccine direct effect against acquisition and progression of the pathogen of interest (VE_*D*_ = 1 − *θ*_*S*_*θ*_*P*_) as *π* and *ω*/*ρ* approach zero, and as *θ*_*S*_ and *θ*_*D*_ approach zero (resulting in near-100% protection). We illustrate the quantitative extent of bias under differing conditions in **Figure 2**.

**Figure 2:**
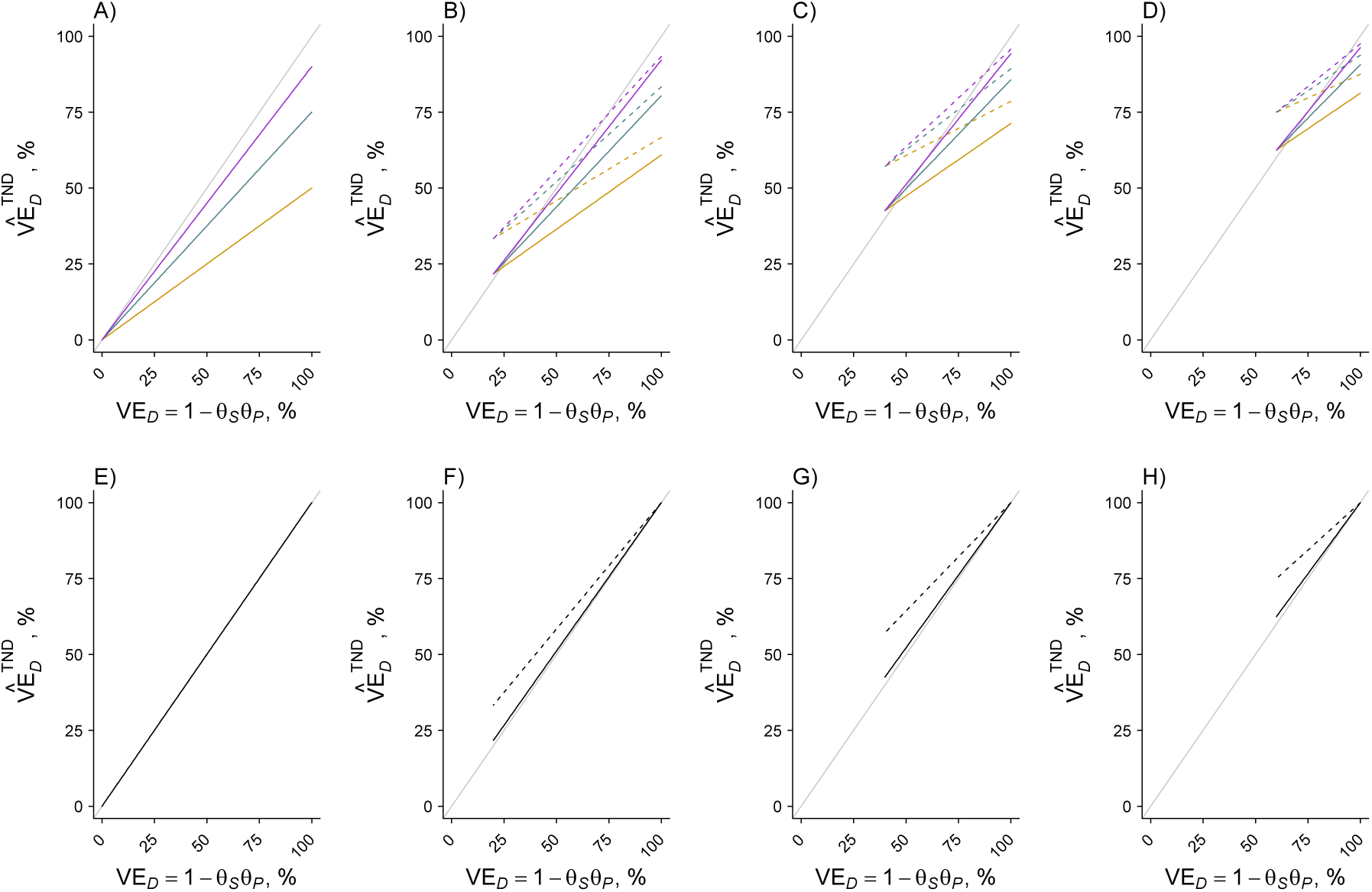
Bias in estimated protection against disease under the test-negative design. We plot the estimated effect of vaccination on risk of disease attributable to the pathogen of interest generated under the test-negative design 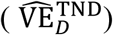, indicating the true vaccine direct effect (VE_*D*_ = 1 − *θ*_*S*_*θ*_*P*_) on the x-axis. Departures from the 1:1 diagonal (grey) line indicate bias. Solid and dashed lines correspond to estimates assuming 10% and 50% prevalence, respectively, of the pathogen of interest among unvaccinated individuals without symptoms (solid, *π* = 0.1; dashed, *π* = 0.5). In the top row (panels A-D), gold, blue, and violet lines correspond to estimates assuming acquisition of the pathogen results in a two-fold, four-fold, and ten-fold increase in individuals’ risk of disease (gold, *ρ*/*ω* = 1; blue, *ρ*/*ω* = 3; violet, *ρ*/*ω* = 9). Organization of panels from left to right corresponds to estimates with increasing vaccine effectiveness against shedding or carriage of the pathogen of interest (VE_*S*_). The left column (panels A and E) assumes 0% protection (*θ*_*S*_ = 1); the second column (panels B and F) assumes 20% protection (*θ*_*S*_ = 0.75); the third column (panels C and G) assumes 40% protection (*θ*_*S*_ = 0.6); and the right column (panels D and H) assumes 60% protection (*θ*_*S*_ = 0.4).

### Bias in the absence of diagnostic uncertainty

We also identify bias under the TND framework under the circumstances addressed in section 7, where detection of the pathogen from cases can be assumed to indicate true disease etiology. Following Eq. 20, in this instance we may consider

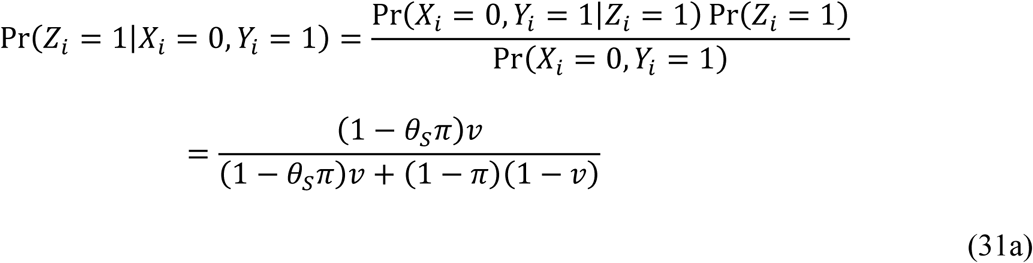

and

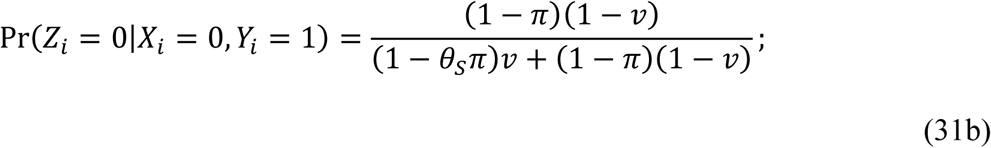

Using the expressions for Pr(*Z*_*i*_ = 1|*X*_*i*_ = 1, *Y*_*i*_ = 1) and Pr(*Z*_*i*_ = 0|*X*_*i*_ = 1, *Y*_*i*_ = 1) presented in Eq. 20, we have

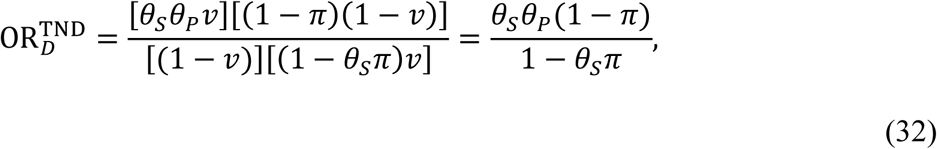

which converges to *θ*_*S*_*θ*_*P*_ as *θ*_*S*_, *θ*_*P*_, or *π* approach zero (**Figure 2**). Otherwise, this approach will overestimate the magnitude of the true vaccine direct effect (i.e., 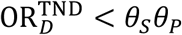).

### Protection against carriage or shedding

Our mathematical framework reveals similar biases affecting estimation of VE_*S*_ under TND-like approaches. Although TND studies typically enroll individuals experiencing a clinical syndrome that could be caused by the pathogen of interest, individuals may, in theory, be matched on any clinical status, including the absence of symptoms potentially attributable to the pathogen of interest. Here, estimates would be expected to indicate vaccine effectiveness against acquisitions that result only in subclinical shedding or carriage (10,33,35).

Consider the odds ratio of vaccination among control individuals who test positive or negative for the pathogen of interest:

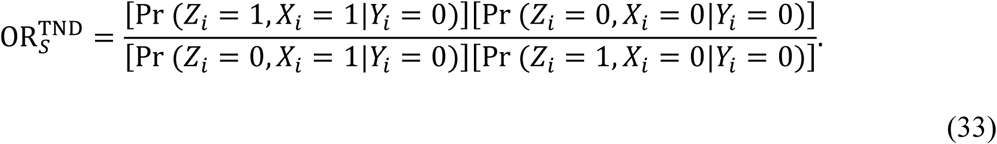

We have presented Pr(*Z*_*i*_ = 1, *X*_*i*_ = 1|*Y*_*i*_ = 0) and Pr(*Z*_*i*_ = 0, *X*_*i*_ = 1|*Y*_*i*_ = 0) in Eq. 4. We may similarly derive

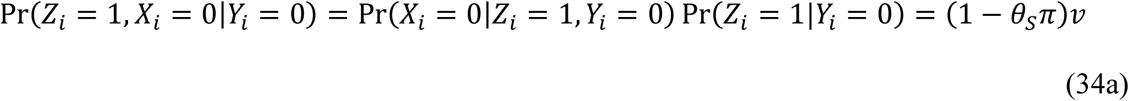

and

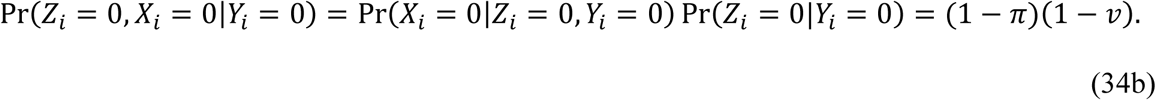

Substituting into Eq. 33, we have

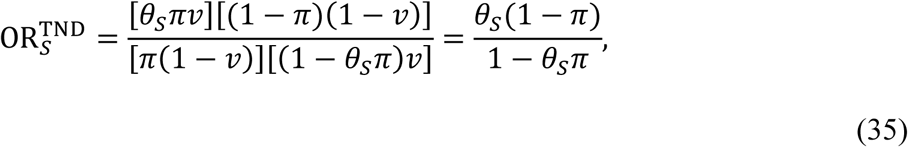

revealing the same form of bias inherent to Eq. 32, above.

## ATTRIBUTABLE FRACTION ESTIMATION

Data from case-control studies such as we describe may also be useful for assessing the fraction of cases attributable to the pathogen of interest. Studies addressing the effects of vaccination on non-specific endpoints provide a framework to “probe” the proportion of cases attributable to the vaccine-preventable pathogen (36,37), based on effects against both all-cause and pathogen-specific disease outcomes.

The OR of vaccination among cases and controls, irrespective of pathogen detection, is

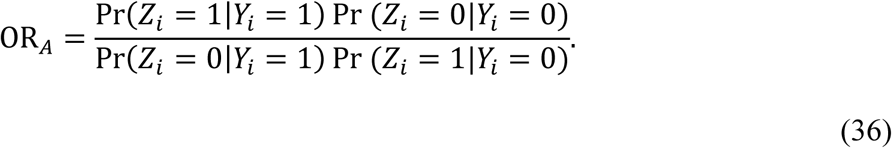

As in Eq. 3, we have Pr(*Z*_*i*_ = 1|*Y*_*i*_ = 0) = *v* and Pr(*Z*_*i*_ = 0|*Y*_*i*_ = 0) = 1 − *v*, while

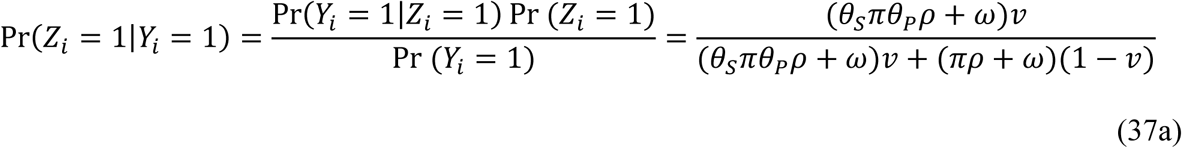

and

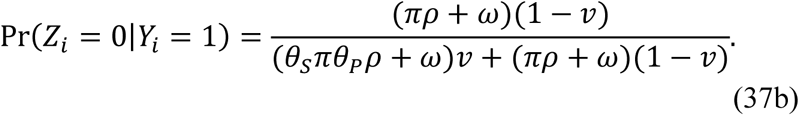

Thus, we may express the effect of vaccination against all-cause disease as

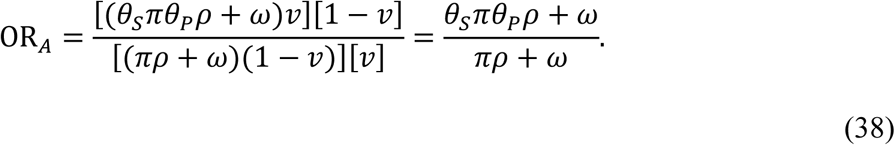

We note that this result equals the risk ratio of disease given vaccination, Pr(*Y*_*i*_ = 1|*Z*_*i*_ = 1) / Pr (*Y*_*i*_ = 1|*Z*_*i*_ = 0).

Rearranging the terms in Eq. 38, we obtain the risk (or rate) ratio of disease attributable to the pathogen of interest, versus other causes, as

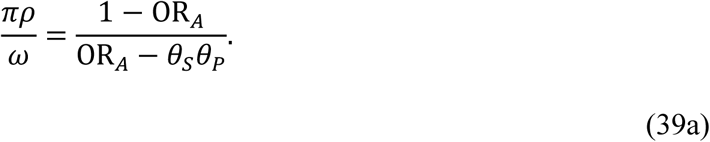

where 1 − VE_*D*_ (Eq. 13) may provide an unbiased input for *θ*_*S*_*θ*_*P*_. Thus, the proportion of disease attributable to the pathogen of interest, absent vaccine-derived protection, is

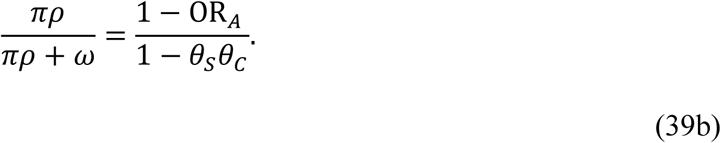

## SUMMARY

Microbiologic diagnosis of certain infectious diseases may be complicated by the prevalence of putative etiologic pathogens among individuals whose symptoms owe to other causes. Here we present a framework for estimating the direct effects of vaccination on different aspects of individuals’ susceptibility through the collection of pathogen detection data in case-control studies. A simple analytic extension provides a “vaccine probe” framework for estimating the proportion of cases attributable to the pathogen of interest.

The approach we have taken also enables assessment of the quantitative extent of bias arising under different scenarios. We identify that pathogen-specific effects of vaccination may be underestimated when a pathogen is detected with low sensitivity among disease cases. Of the disease-specific applications we have described, this circumstance likely has the greatest importance for pneumonia, where pathogens may continue causing disease in the lung after clearance from their commensal niche in the upper respiratory tract. This circumstance would presumably reduce sensitivity of pathogen detection among disease cases to a greater extent than what is expected among controls.

We also identify that TND studies may produce biased vaccine effectiveness estimates either with or without misclassification of symptomatic individuals based on pathogen detection. Notably, we identify that bias in TND studies may result in either over-estimation or under-estimation of true vaccine effectiveness, making the interpretation of estimates difficult. Here we do not consider additional issues that have been found to impact the reliability of estimates from TND studies, including individuals’ acquisition of immunity following natural infection and the mode of vaccine action (35,38). Nonetheless, our findings add to a growing list of concerns about uses of the TND for estimation of vaccine effectiveness (10,14,39). Control groups without symptoms potentially attributable to the pathogen of interest may be of value to correct for biases arising in conventional TND studies.

Case-control studies are commonly undertaken to estimate the direct effects of vaccination. Collecting pathogen detection data from controls, alongside symptomatic cases, presents a simple strategy to correct for misclassification of symptoms not attributable to a detected pathogen. Provided such data are collected, the approaches we describe permit estimation of vaccine effectiveness against pathogen-specific endpoints without a diagnostic gold standard.

## Data Availability

No data are presented in this manuscript

## Abbreviations

OR: Odds ratio
TND: Test-negative design
VE: Vaccine effectiveness

